# Engaging doctoral students in peer review: a pre-post study evaluating the effectiveness of the “Peerspectives” course on review quality, knowledge and skills

**DOI:** 10.1101/2025.02.11.25322060

**Authors:** Jessica L. Rohmann, Nadja Wülk, Kerstin Rubarth, Hannah Grillmaier, Iman Abdikarim, Mariana Lopes Simoes, Sara Schroter, Marco Piccininni, Tobias Kurth, Toivo Glatz

## Abstract

**Objective:** To assess the effectiveness of the *Peerspectives* course on editor-judged quality of peer review

**Design:** Single arm, pre-post interventional study

**Setting:** *Peerspectives* is a peer review training course developed for doctoral students at the Charité – Universitätsmedizin Berlin, Germany

**Participants:** Doctoral students in health research domains who enrolled in the *Peerspectives* course between October 2020 and August 2022 and consented to participate (*N* = 80). **Intervention:** A semester-long course (approx. 18 weeks) providing training about the structure, purpose, and conduct of peer review and editing processes in biomedical journals. The course consisted of 12 hours of interactive lectures, weekly homework assignments, and 12 hours of hands-on, small-group workshops under guidance of mentors with reviewing and editing experience. Students peer-reviewed real scientific manuscripts under review at a partnering journal, *The BMJ*.

**Main outcome measures:** The overall quality of the peer review reports as judged by two independent *BMJ* editors using the global score of the Review Quality Instrument (RQI) pre- and post-intervention. Additionally, participants’ self-assessed knowledge and skills related to scholarly peer review (1-5 Likert scale).

**Results:** After course completion, participants’ RQI scores were higher than before the course (median increase of 0.5 points, p<0.001; mean increase of 0.36 points, p<0.001). The RQI scores of participants’ post-course reports were not non-inferior to those of actual *BMJ* reviewers for the same manuscripts. Self-assessed peer review-related knowledge skills increased across all questionnaire items after course completion. Largest improvements were seen in understanding what is expected from a reviewer (increase in means from 2.9 to 4.5), confidence in own ability to review (2.5 to 3.9), and knowing what to look for in a manuscript while reviewing (2.8 to 4.2).

**Conclusions:** Providing doctoral students with comprehensive training resulted in an editorially significant increase in review report quality and improved understanding of the role and expectations of peer reviewers in the scholarly publishing processes and confidence in giving constructive feedback.

**Study pre-registration:** https://osf.io/vndcx

## INTRODUCTION

Scientific journals publish scholarly articles, providing an important platform for transparent presentation, exchange, and discussion of new scientific developments. Peer review remains a backbone of scholarly publishing, though often criticized for being ineffective, unfair, and unreliable.[1–6] Critics disagree about which aspects of today’s peer review are most problematic and how to best ensure its relevance, integrity and quality, and how to improve its sustainability.[2,7–9] In the 2018 Publons Global State of Peer Review report, 88% of survey respondents indicated that training is important or extremely important for ensuring high-quality peer review.[10] Despite expressing great interest in receiving further training,[11–13] many scientists report lacking guidance on how to review a scientific paper.[13,14]

Because of the steady increase in the number of submitted articles, coupled with limited rewards or incentives for peer review, reviewers experience an increasing workload and growing reviewing fatigue.[5,6,10,15] Nevertheless, scientists invest considerable amounts of effort and time into the publication system; peer review activities have been estimated to cost more than 100 million researcher hours globally per year, with the time contributed by US-based reviewers alone valued at over $1.5 billion USD and UK-based reviewers nearly $400 million USD in 2020.[16] Simultaneously, journal editors report difficulties finding reviewers able to reliably deliver high-quality peer review reports.[10,17,18] This leads to an increased editorial workload and publishing delays – challenging conditions only exacerbated during the COVID-19 pandemic.[6,19,20]

Given their importance, aspects of scholarly publishing, including peer review, remain surprisingly absent in the curricula of advanced academic programs in the health sciences. At the same time, early career researchers (ECRs) represent a talent pool with great interest in improving research culture and practice.[21] Their recent coursework may provide them with familiarity with the latest scientific developments and methods, and they may also have more time to dedicate to peer review activities compared to senior scientists. Peer review training, if any, has traditionally followed an informal apprenticeship model.[22] Existing peer review training resources for biomedical researchers[23] generally do not involve the review of actual manuscripts under consideration for publication, and most are developed by publishers,[24–28] who seem to be targeting active reviewers and more experienced scientists. Whether prospective peer reviewers make use of these tools and whether they are effective remains unknown.[22] We identified only a few efforts to explicitly engage students in the peer review of scientific articles.[29,30] The limited existing evidence indicates that short-duration training,[31] receiving written feedback from editors,[32] or the simple pairing of novice and experienced reviewers[33,34] are of limited value in improving review-related knowledge or review quality.

We developed *Peerspectives* to bridge the aforementioned gaps. This semester-long, researcher-led peer review training initiative combines interactive lectures with small-group workshops led by experienced mentors with editing and peer review experience. By using actual scientific manuscripts under review at a partnering journal, *The BMJ*, *Peerspectives* sought to enable doctoral students to make tangible contributions to biomedical research while learning about peer review and the scientific publishing landscape.

The present study aimed to determine whether and to what extent the completion of the Peerspectives curriculum had an effect on editor-assessed quality of peer review reports drafted by doctoral students. As secondary aims, we investigated whether the quality of the review reports drafted by *Peerspectives* participants after course completion were non-inferior to those submitted by actual *BMJ* reviewers, and we assessed differences in self-assessed knowledge and relevant peer review competencies among participants following course participation.

## METHODS

### Study design

We conducted a single arm, pre-post study.[35] All study participants received the intervention, and the outcome was assessed twice: once before and once after the intervention was administered. The differences between the two outcome measurements across the individuals quantified the average effect of the intervention at the end of the study.[36] The key assumption of this analysis is that if study participants had not received the intervention, their second outcome measurement would have been the same as their first outcome measurement.[36] In more technical terms, pre-post comparisons rely implicitly on a stationarity assumption of the untreated counterfactuals — that is, the outcomes that would have been measured if no intervention had been implemented.[36,37] Despite requiring stronger assumptions than randomized studies,[35,36] pre-post designs are common in studies assessing the effect of educational interventions.[36]

### Participants

All students enrolling in the *Peerspectives* course in the semesters October 2020 - March 2021, April 2021 - August 2021, October 2021 - March 2022, and April 2022 - August 2022 were invited to participate in this scientific evaluation study, which was approved by the ethics committee of the Charité – Universitätsmedizin Berlin (EA4/190/20).

After reading the detailed participant information materials, interested students were asked to provide written informed consent. Study participation was not required to enroll in the course, and all students were informed that their choice not to participate in the study would not impact their ability to pass the course nor receive course credit. Instructors were unaware of which students had elected to participate in the evaluation study.

Course enrollment capacity was contingent on the availability of workshop group mentors and fluctuated between 16 and 23 students per semester. Health Data Sciences PhD Program students at the Charité – Universitätsmedizin Berlin were prioritized for course enrollment. We opened additional spots to other doctoral students both within and outside of our institution and advertised the course via our institute’s website, on social media, and via our instructors, editor-mentors, PhD program coordinators, and course alumni.

To confirm participation in the course, selected students were required to sign a confidentiality agreement in which they agreed not to disclose information to third parties about the unpublished manuscripts encountered during the training and assessment activities and made available by *The BMJ*, the partnering journal. Consenting study participants were additionally asked to provide information about their age, gender, educational background (highest degree, anticipated doctorate completion date), any prior epidemiology or statistics training, and prior reviewing experience on a short questionnaire sent via email prior to the start of the course (**Supplementary Material A**).

### Intervention

The *Peerspectives* course ran over approximately 18 consecutive weeks (slightly variable due to public holidays) and was designed to provide insights into the structure, purpose, and conduct of the peer review and related editing processes in biomedical journals. We provide additional details on the course structure and format (**Box 1**) and a sample syllabus (**Supplementary Material B**). The language of instruction was English, and the course was offered free of charge. All meetings were held online using Zoom (Zoom Video Communications Inc.) initially due to the COVID-19 pandemic and later in an effort to reach students and engage editor-mentors located outside of Berlin.

## Box 1. Short description of the *Peerspectives* peer review training course

The first part of the course consisted of four 3-hour interactive lectures led by instructors affiliated with the Health Data Sciences PhD Program at the Charité – Universitätsmedizin Berlin. These lectures focused on: (1) the role of scientific journals, editors, peer reviewers, and authors in scientific publishing; (2) ethical guidelines for peer review, open science, and reporting of sex and gender-related aspects; (3) the conduct of peer review, including step-by-step guidance on how to write a constructive peer review report; and (4) a live demonstration of how to draft a peer review report, including writing tips, for an actual “live” manuscript under review. After each lecture, students were assigned homework to be submitted prior to the next session, which started with a group discussion of take-home tasks.

In the second part of the course, groups of four (occasionally five) students were assigned to work together in small groups under the guidance of an experienced editor-mentor to produce peer review reports for four “live” manuscripts per group contemporaneously under review at *The BMJ*. These editor-mentors donated their time to the project and did not receive remuneration. They were not involved in the editorial handling of the manuscripts, data collection, or analysis. Each workshop group produced four review reports, totaling 16 to 20 review reports per course run.

In the week prior to each of the four workshops, a PDF file of the manuscript and any supplemental materials were uploaded to a secure channel on PaperHive (PaperHive UG, Berlin, Germany), a protected web platform. This was accessible by all group members, allowing collaborative annotation of the manuscript and real-time exchange of comments.[38] The students were instructed to create a first draft of each peer review report together, with one student taking a lead role on a rotating basis. During the 3-hour workshop, this draft was discussed and revised with the editor-mentor.

Once the peer review report was finalized and approved by all students and the group’s editor-mentor, the editor-mentor submitted it to *The BMJ,* including the names of all students as contributors, which were visible to the authors of the submitted papers and editors (and readers, if ultimately published) as part of *The BMJ*’s open peer review process. Once available, the editor-mentors were instructed to disseminate and discuss the other reviewers’ comments and the final editorial decision on the paper with their group members.

All students (regardless of study participation) were required to attend all lectures and workshops, submit all homework assignments, actively prepare for and contribute to the workshops, and complete the pre- and post-course peer review report assessments to receive four European Credit Transfer and Accumulation System (ECTS) credit points.[39] To compensate for missed sessions, we provided make-up assignments. To wrap up the course, we offered an optional final session, in which the course participants could listen in on a *BMJ* editorial manuscript meeting, followed by a group reflection with course instructors. Upon completion, all students were invited to complete an anonymous course evaluation on LimeSurvey (LimeSurvey GmbH, Hamburg, Germany) to provide feedback to instructors on course content, mode of delivery, and organization. See **Supplementary Material B** for a sample course syllabus.

### Outcomes

#### Primary outcome

All study participants were required to draft two peer review reports on their own for two scientific manuscripts: one assigned to them before the course commenced (“pre-intervention”) and one immediately after completing the final workshop session of the course (“post-intervention”). Students were explicitly instructed to work on these assessments alone and not discuss them with others; however, they were told they could freely use any other resources (“open-book”).

To simulate real-world reviewer conditions, students received complete instructions using a standardized email template akin to the peer review invitations sent by *The BMJ*. Like many major biomedical journals, we provided students with a two-week deadline to complete their peer review assignments. Reminders mimicking the manuscript management platforms’ chasing mechanisms were sent to those who had not submitted their reports one week before, one day before, and again on the day of the deadline. Participants could request a one-week extension; in these cases, we sent additional reminders at the same intervals. In cases of non-responsive participants, we sent additional reminders until overdue review reports were received.

For these individual assessments, a set of four manuscripts were selected from those contemporaneously under review at *The BMJ* one month prior to the commencement of each course run. Each semester, we block randomized participants into four groups using the *randomizr* R package[40] v0.24.0. One manuscript was assigned at random to all members of each group for the pre-intervention assessment and a different one for the post-intervention assessment.

The primary outcome was the editor-judged overall quality of the study participants’ peer review reports. Each report was scored using the eighth item (*How would you rate the quality of this review overall?*) of the Review Quality Instrument (RQI) Version 3.2 on a five-point Likert scale ranging from 1 - “poor” to 5 - “excellent”.[41] This validated instrument was specifically developed to assess review quality and has been used in several previous studies.[31,42]

The same two *BMJ* editors scored all peer review reports for a given manuscript: the manuscript’s actual handling editor and one additional editor, none of whom was an instructor in the course. The pseudonymized reports were scored in an online form created in LimeSurvey (LimeSurvey GmbH, Hamburg, Germany), and the graders were blinded to whether the report had been written pre- or post-intervention. To mitigate potential order effects, we presented raters with their scoring assignments in a random order. All raters were experienced in using the RQI global item of review quality, as it is routinely used to score all peer review reports received by *The BMJ*. The primary outcome was defined as the mean RQI global score given by the two raters to each participant.

#### Secondary outcomes

For a secondary analysis, the routinely collected global review quality score for the actual peer review reports received by *The BMJ* were extracted from the ScholarOne journal management system, which had been scored by the manuscript’s handling editor. This analysis was not included in the original study pre-registration.

Before and after the intervention, we also collected information about self-assessed knowledge and skills related to scholarly peer review. Prior to course commencement, all study participants were asked to complete an online survey containing eight questions to self-assess perceived confidence in specific peer review-related knowledge and abilities, e.g., “I feel confident in formulating constructive and clear scientific critique” (**Supplementary Material C**). The structure of this pre-intervention survey was based on the Personality Evaluation Inventory (PEI), designed to assess perceived confidence across various domains relevant to college students.[43] Response options were on a 5-point Likert scale (1 - “strongly disagree” to 5 -“strongly agree”). The same survey was administered again upon submission of the post-intervention peer review report. Students had one month to complete this final self-assessment and were reminded up to three times.

#### Statistical Analysis

We performed analyses according to our a priori statistical analysis plan (https://osf.io/vndcx), which included a simulation-based sample size calculation using expected distributions of the pre- and post-intervention primary outcome. The secondary comparison of the global RQI scores with actual *BMJ* reviewers was added after the protocol was created. We present descriptive statistics, including medians, interquartile range limits, means, and standard deviations.

In the primary analysis, we assessed the effect of the intervention, taking part in the *Peerspectives* course, on peer review report quality at the time of the second outcome assessment, among study participants. To do this, we compared the pre- and post-intervention primary outcome scores (mean global RQI) of the study participants. Given the paired nature of the measurements, we used a non-exact asymptotic paired Wilcoxon signed-rank test and a paired t-test to determine whether the intervention had a statistically significant effect on the primary outcome. Both tests were two-sided, with a significance threshold of 0.05.

Next, we investigated whether the participants’ post-intervention RQI scores were non-inferior to those of the actual *BMJ* peer reviewers assigned to the same manuscript. We compared the handling editor’s global RQI scores for actual BMJ reviewers with the same editor’s global RQI scores for the *Peerspectives* course participants. We employed a nonparametric statistical approach that accommodates the clustering arising from the interdependence of ratings within manuscripts.[44] This method infers the ‘relative effect’, also known as the generalized Wilcoxon-Mann-Whitney effect.[45]

A 0.4 unit difference in the average total score of the first seven RQI items was considered “editorially significant” in prior work,[31] so we made the assumption that a 0.4 difference on the global item would also be "editorially significant”. To express this statement in terms of a relative effect, the gold standard RQI scores from the actual *BMJ* reviewers were utilized to create a dataset of hypothetical *Peerspectives* participant scores. This was done by randomly splitting the gold standard RQI data into two parts: 60% of the scores remained unchanged, while for the remaining 40%, scores were reduced by 1 point. The resulting mean difference between the two datasets was 0.4, corresponding to the threshold of a difference that would be considered “editorially significant”. We defined the non-inferiority null hypothesis as a relative effect ≤ 0.379, corresponding to the relative effect in this hypothetical simulated data.

As a further secondary analysis, we tested for differences in self-assessed knowledge and relevant peer review competencies between pre- and post-intervention measurements using a paired Wilcoxon signed-rank test and a paired t-test, as described above. Since we made multiple comparisons in this analysis, we corrected all p-values for multiple testing using Bonferroni’s method (adjusted for 8 comparisons).

Owing to the very low number of missing values, complete case analyses were performed for the primary and secondary analyses. We performed two supplementary analyses. First, as a sensitivity analysis, we excluded those participants who did not turn in their final self-assessment of knowledge and skills questionnaire. Second, we report results stratified by prior peer review experience.

Finally, we computed inter-rater agreement using the weighted Kappa for the review quality ratings by handling vs. non-handling editors using the R package *psych*[46] version 2.3.3.

All statistical analyses were performed using R version 4.4.2.

#### Patient and public involvement

The *Peerspectives* program and this linked scientific study were developed for doctoral students, with considerable input from early career researchers (the target population) at all stages. Early career researchers were heavily involved in the curriculum development, course instruction and organization. After completion of the course, participants provided feedback, which was used to improve the content and logistics for future course runs. We also involved doctoral students, instructors, and journal editors in conducting this study. The quality metric for assessing peer-review reports was chosen in consultation with *BMJ* editors.

## RESULTS

Over four semesters, 82 doctoral students enrolled in the *Peerspectives* course, of which 80 provided written informed consent to participate in the scientific evaluation study (see flowchart, **Figure 1**). Upon enrollment, the study participants had a mean age of 30 years and 59% reported being female (see **Table 1**). The majority of participants reported their highest level of completed education as a Master’s degree (75%), while the remainder reported studying medicine. Approximately one-third of students reported having prior peer reviewing experience, and those with prior reviewing experience reported having reviewed a median of two articles (IQRL: 1.0 - 3.0). Two participants dropped out of the study before any post-intervention assessment. The average RQI global scores for the pre- and post-intervention assessments were available for the remaining 78 participants. Of these, six did not submit the post-intervention self-assessment questionnaire. Associations between the pre-intervention primary outcome and age or number of prior peer reviews are presented in **Supplementary Material D**.

**Figure 1.**
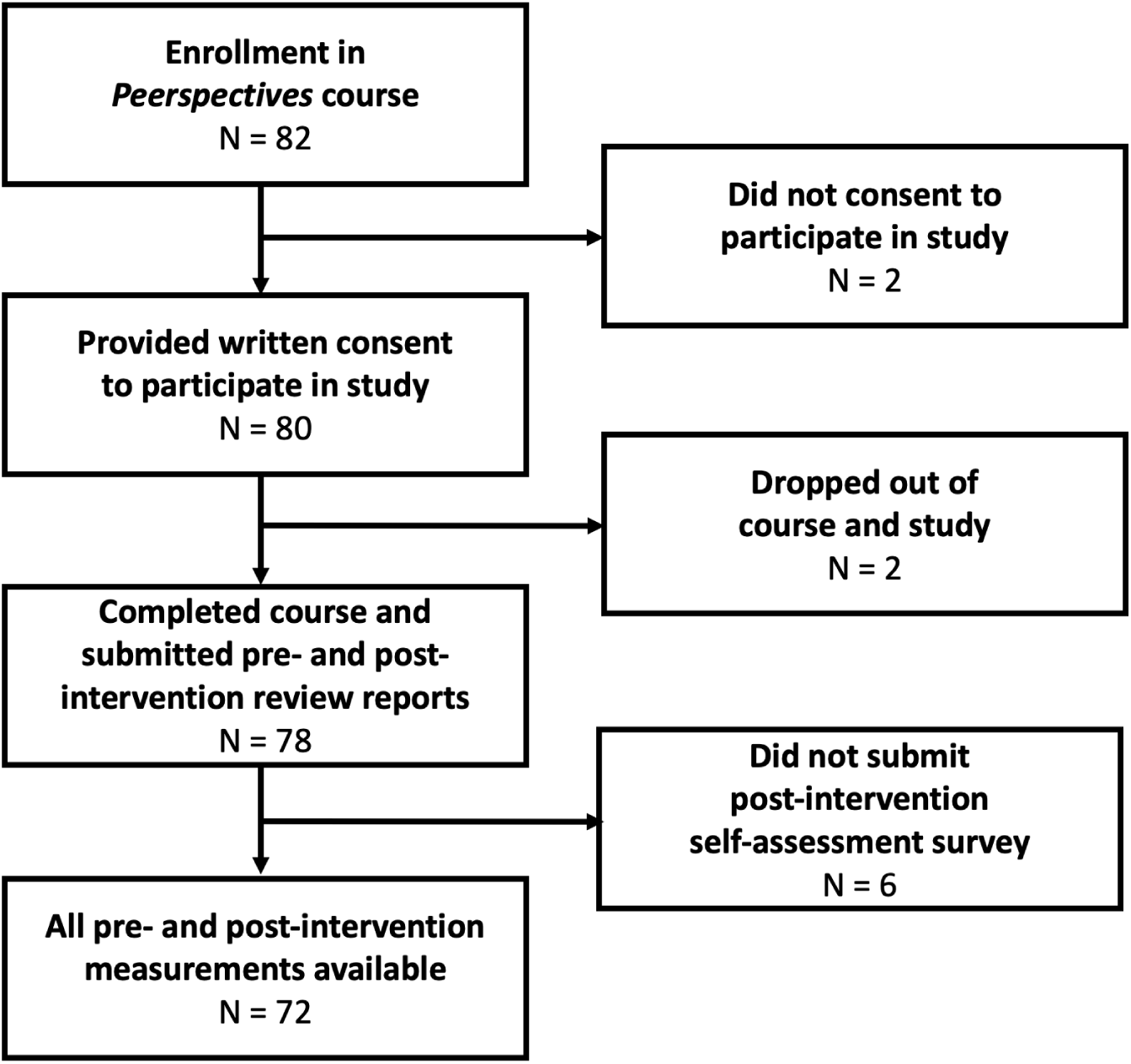
Participant flowchart.

**Table 1.** Participant characteristics before the intervention.

|  |  | Overall (N = 80) |
| --- | --- | --- |
| **Age** | Mean (SD) | 30.3 (5.1) |
|  | Range | 22 - 49 |
| **Gender** | Female | 47 (59%) |
|  | Male | 31 (39%) |
|  | Other | 0 (0%) |
|  | Prefer not to say | 2 (2%) |
| **Highest degree** | Master | 60 (75%) |
|  | Medicine | 20 (25%) |
|  |  | **With prior peer review experience (N = 26)** |
| **Number of prior peer reviews performed** | Median (IQRL) | 2.0 (1.0 - 3.0) |
|  | Range | 1 - 15 |

*Abbreviations:* SD: standard deviation, IQRL: interquartile range limits

In the primary analysis, we compared the quality of the peer review reports before and after participation in *Peerspectives*. In total, 46 participants had a higher primary outcome on the post-intervention review report, 18 remained the same, and 14 were worse (see **Figure 2**). Prior to participating in the course, the median primary outcome was 3.5 (IQRL: 3.0 - 4.0); after the course, the median was 4.0 (IQRL: 3.5 - 4.0). We found a statistically significant difference between pre- and post-intervention primary outcome (*p* < .001). The mean primary outcome also increased, from 3.47 (*SD* = 0.72) before the intervention to 3.83 (*SD* = 0.59) after the intervention (mean difference: 0.36, *t* = -4.35, *p* < .001).

**Figure 2.**
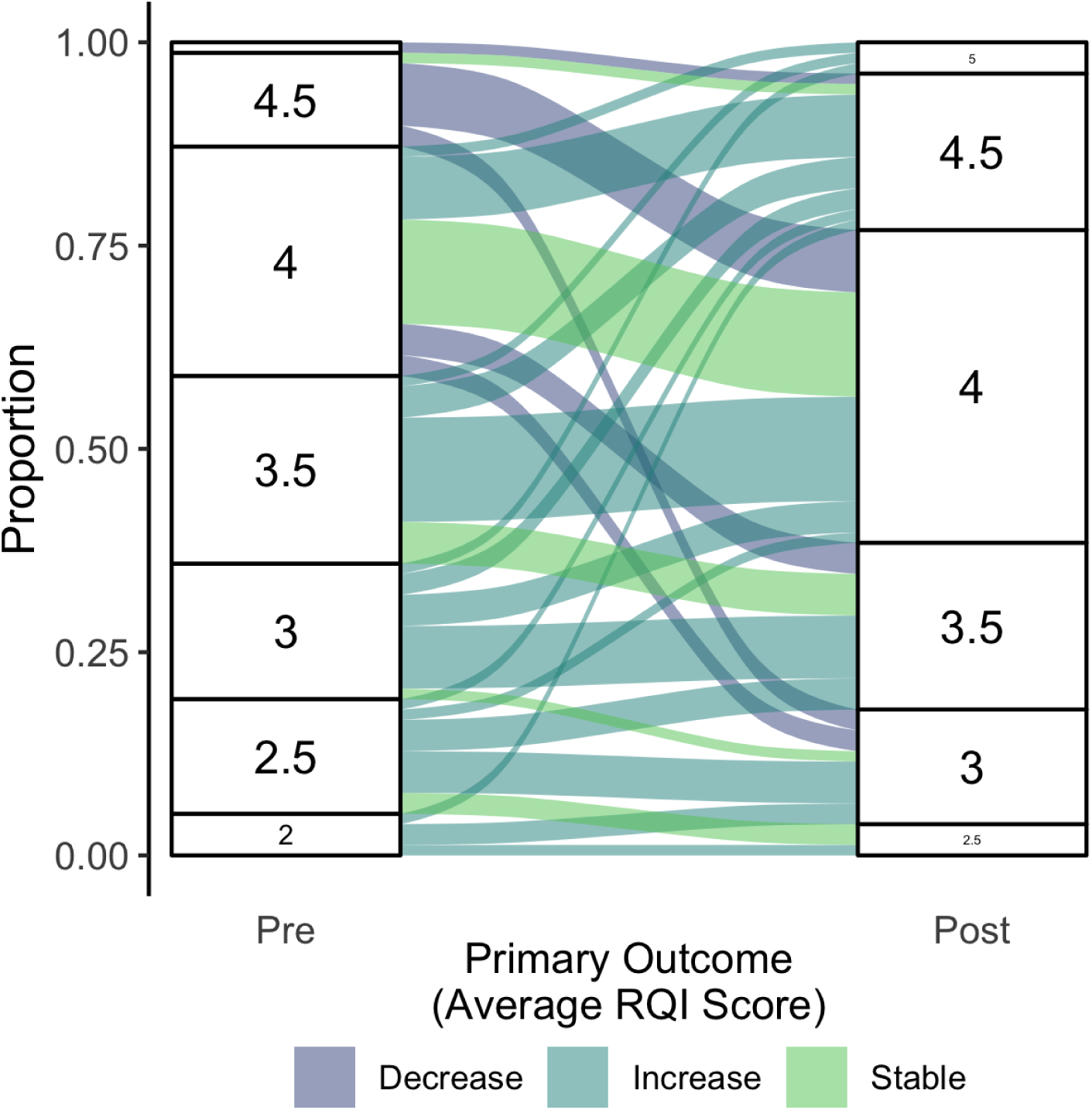
Alluvial diagram showing changes in the primary outcome (average global RQI scores of ratings by two BMJ editors) between pre- and post-intervention. RQI scores range from 1 to 5, with 5 indicating better quality. RQI: Review Quality Instrument.

*Abbreviations: SD*: standard deviation, IQRL: interquartile range limits

Compared to the Peerspectives participants, *BMJ* reviewers had a higher proportion of excellent (“5”) RQI global scores (30% vs. 6%), as assessed by each article’s handling editor. Therefore, the actual *BMJ* reviewers had an overall more favorable score distribution than the *Peerspectives* participants. However, all Peerspectives participants received a score of 3 or higher, whereas 4% of *BMJ* reviewers delivered reports scored with 2; no report received a poor (“1”) score (see **Figure 3**). For each manuscript used in the assessment, the median RQI scores of participants post-intervention and the median scores obtained by *BMJ* reviewers are shown in **Supplementary Material E**.

**Figure 3.**
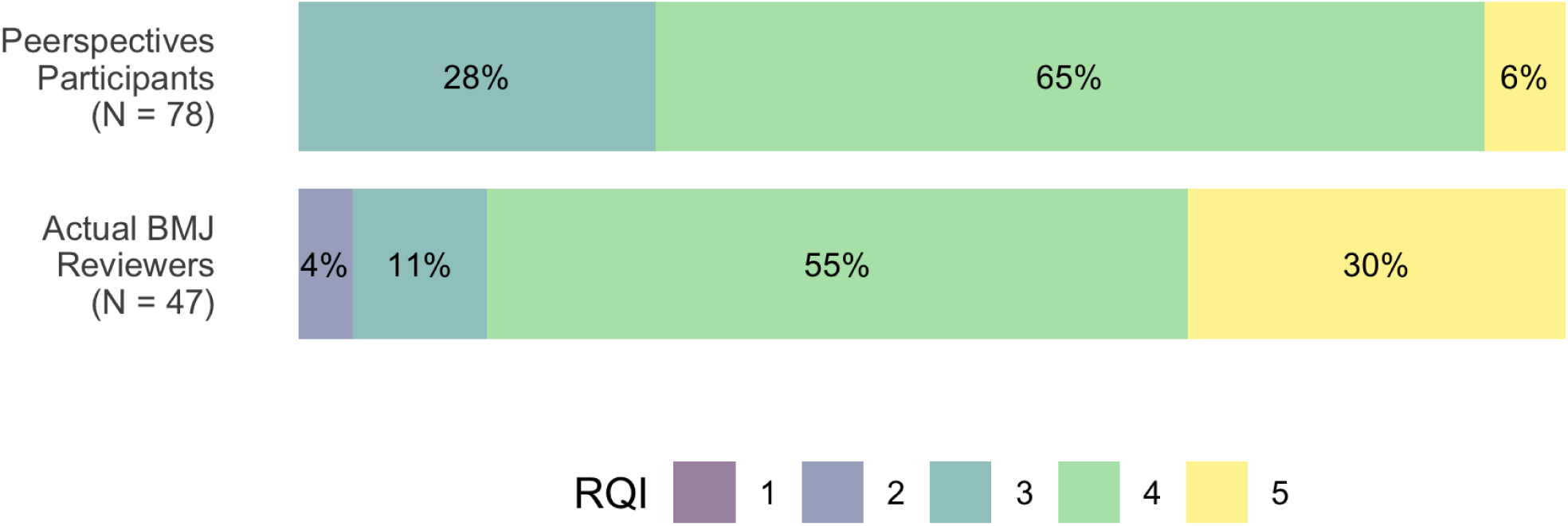
Distribution of global RQI scores ranging from 1 (poor) to 5 (excellent) for Peerspectives participants after course completion and actual BMJ reviewers, as rated by the handling editor. Percentages may not add up to 100% due to rounding. RQI: Review Quality Instrument

In the secondary analysis, we obtained an estimate for the relative effect of 0.36. When testing the null hypothesis of the non-inferiority test, we obtained a *p*-value of 0.655. Thus, we were unable to demonstrate non-inferiority of the *Peerspectives* participants’ RQI global scores compared to the actual *BMJ* reviewers’.

Finally, upon comparing the pre- and post-intervention self-assessments of peer review-related knowledge and skills, we observed statistically significant improvements in all questionnaire items (all Bonferroni adjusted *p*-values < .001). The individual participant trajectories are shown in **Figure 4** and Bonferroni adjusted *p*-values from statistical tests are summarized in **Table 2**. Notably, the categories 4 - “agree” and 5 - “strongly agree” greatly dominated the responses in the post-intervention assessments for all questionnaire items in contrast to the pre-intervention assessments, in which the three middle categories 4 - “agree”, 3 - “neither agree nor disagree”, and 2 - “disagree” were approximately evenly distributed.

**Figure 4.**
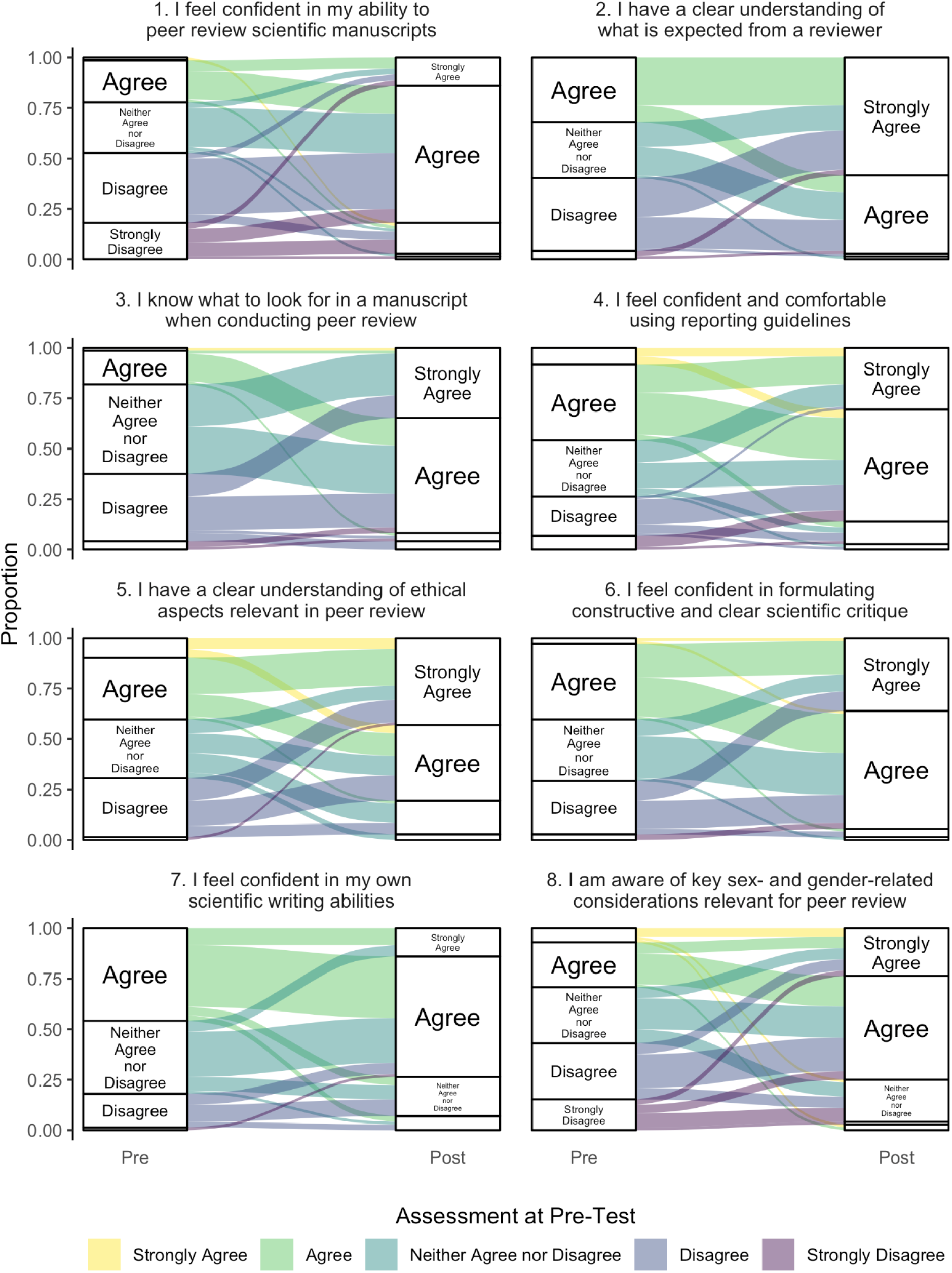
Alluvial diagram showing changes in individuals’ self-assessed peer review-related knowledge and skills between pre- and post-intervention assessments.

**Table 2.** Participants’ self-assessed peer review-related knowledge and skills before and after intervention.

| **Statement (N = 72)**<br>(1 = strongly disagree,<br>5 = strongly agree) | **Pre-**<br>**assessment**<br>mean (SD)<br>median (IQR) | **Post-**<br>**assessment**<br>mean (SD)<br>median (IQR) | **Wilcoxon**<br>**signed-**<br>**rank test**<br>**p-value\*** | **Paired**<br>**t-test**<br>**p-value\*** |
| --- | --- | --- | --- | --- |
| 1. I feel confident in my ability to peer review scientific manuscripts | 2.5 (1.1)<br>2.0 (2.0 - 3.0) | 3.9 (0.7)<br>4.0 (4.0 - 4.0) | < .001 | < .001 |
| 2. I have a clear understanding of what is expected from a reviewer | 2.9 (0.9)<br>3.0 (2.0 - 4.0) | 4.5 (0.6)<br>5.0 (4.0 - 5.0) | < .001 | < .001 |
| 3. I know what to look for in a manuscript when conducting a peer review | 2.8 (0.8)<br>3.0 (2.0 - 3.0) | 4.2 (0.7)<br>4.0 (4.0 - 5.0) | < .001 | < .001 |
| 4. I feel confident and comfortable using reporting guidelines | 3.2 (1.1)<br>3.0 (2.0 - 4.0) | 4.1 (0.7)<br>4.0 (4.0 - 5.0) | < .001 | < .001 |
| 5. I have a clear understanding of ethical aspects relevant in peer review | 3.2 (1.0)<br>3.0 (2.0 - 4.0) | 4.2 (0.8)<br>4.0 (4.0 - 5.0) | < .001 | < .001 |
| 6. I feel confident in formulating constructive and clear scientific critique | 3.1 (0.9)<br>3.0 (2.0 - 4.0) | 4.3 (0.7)<br>4.0 (4.0 - 5.0) | < .001 | < .001 |
| 7. I feel confident in my own scientific writing abilities | 3.3 (0.8)<br>3.0 (3.0 - 4.0) | 3.8 (0.8)<br>4.0 (3.0 - 4.0) | < .001 | < .001 |
| 8. I am aware of key sex- and gender-related considerations relevant for peer review | 2.8 (1.2)<br>3.0 (2.0 - 4.0) | 3.9 (0.9)<br>4.0 (3.8 - 4.0) | < .001 | < .001 |

\* *p*-values adjusted for 8 comparisons using Bonferroni's method. IQR: interquartile range limits, SD: standard deviation

The largest improvements were observed in having a clear understanding of what is expected from a reviewer, with an increase in means from 2.9 to 4.5, the confidence in one’s own ability to peer review scientific manuscripts (2.5 to 3.9), and for knowing what to look for in a manuscript when conducting a peer review (2.8 to 4.2).

### Supplementary analyses

After excluding those participants who did not submit their post-intervention self-assessment of knowledge and skills questionnaire (*N* = 6), we obtained identical results to the primary analysis. After stratifying by self-reported prior peer review experience, those who had never reviewed before (*N* = 53) had a median primary outcome of 3.5 (IQRL: 3.0 - 4.0) pre-intervention and 4.0 (IQRL: 3.5 - 4.0) post-intervention. Comparing the mean primary outcome, we observed a 0.43 point increase (pre-intervention: 3.42, *SD* = 0.76; post-intervention: 3.85, *SD* = 0.60) among those with no prior reviewing experience. Both Wilcoxon signed-rank test (*p* < .001) and paired t-test were statistically significant (*p* < .001). Those who had reviewed before (*N* = 25) had a median primary outcome of 3.5 (IQRL: 3.0 - 4.0) before the intervention and 4.0 (IQRL: 3.5 - 4.0) after completion of the course. The group with prior reviewing experience saw a smaller increase in mean primary outcome of 0.22 points between pre- (3.58, *SD* = 0.69) and post-intervention (3.80, *SD* = 0.56). Still, both Wilcoxon signed-rank test (*p* = .043) and paired t-test (*p* = .038) were statistically significant.

#### Inter-rater agreement

In the primary analysis, since each individual’s review report for a given manuscript was scored by two out of four *BMJ* editor raters, three possible pairings of editors were possible for which we could calculate inter-rater agreement. With sample sizes of 74, 40, and 42 reports scored by pairs of editors, the weighted Kappa values were 0.53, 0.50, and 0.49, respectively, indicating a “moderate” inter-rater agreement in the RQI scoring.

## DISCUSSION

Following participation in the *Peerspectives* peer review training course, we observed an improvement in the overall quality of participants’ peer review reports. The average RQI global score rated by two blinded journal editors was statistically significantly higher after the course, with a 0.36 increase in the mean and a 0.5 increase in the median. Given that prior work considered thresholds of a 0.4 unit difference[31] and a 10% difference in RQI scores[47] as “editorially significant”, the improvement we observed in the primary outcome appears meaningful. Our supplementary analysis showed that the quality of review reports of those participants with some prior reviewing experience and those with none both improved with the intervention.

Also, the participants’ own assessments of their knowledge and skills relevant for peer review improved. Largest improvements were seen in understanding the expectations in the role of a peer reviewer, knowing what to look for in a manuscript when reviewing, and confidence in conducting peer review. Following the course, participants reported feeling more confident in expressing scientific critique, using reporting guidelines, and in their own scientific writing abilities. These acquired skills may make them more likely to accept future review assignments and follow through in delivering high quality peer review reports useful to both authors and editors.

Despite these improvements, we could not demonstrate non-inferiority of the overall quality of the participants’ post-intervention peer review reports compared to those of the actual *BMJ* reviewers who had been invited to review the same manuscripts. This finding is not unexpected considering *BMJ* reviewers represent a highly-selected group, many of whom likely reviewed for *The BMJ* in the past and achieved sufficiently high quality scores to be invited to review again. Still, 71% of *Peerspectives* participants received RQI global scores of 4 or 5 (out of 5) by the handling editor, which is noteworthy given that the participants were all doctoral students.

*Peerspectives* was originally conceptualized to teach peer review to doctoral students in the biomedical sciences domains in Germany, a setting in which doctoral students are increasingly pursuing cumulative, publication-based dissertation projects.[48] ECRs are notably absent in peer reviewing and editing positions and, therefore, not generally the target of peer review training initiatives despite representing a unique and highly relevant group seeking formal training as peer reviewers. Our results show that ECRs are keen to be trained, and can draft good quality peer review reports even at a very early stage given sufficient guidance and supervision. As it remains a challenge for journal editors to find peer reviewers willing and able to deliver informative, high-quality peer review reports under tight deadlines,[6,49] engaging ECRs in peer reviews may represent a benefit for both parties: ECRs and scientific journals.

ECRs who are drafting their first scientific manuscripts may benefit from learning what journal editors and peer reviewers look for, and gain firsthand insights into an unfamiliar assessment process that is often used as external quality control and often relevant (either directly or indirectly) for their academic qualification. By reviewing actual manuscripts in the hands-on workshops and having the opportunity to observe a *BMJ* editorial meeting, *Peerspectives* participants were directly exposed to editorial decision-making processes. Not only could they receive credit for their doctoral studies but they were also credited by name for their reviewing work in *The BMJ’s* open peer review procedure. After course completion, we received overwhelmingly positive course evaluations. Though not formally analyzed for this publication, the responses highlight identified strengths of the course content, format and delivery. We provide a selection of free text responses to the question of whether participants would recommend the course to others and if so, to whom (**Supplementary Material F**).

Students may not be the only ones to benefit from comprehensive peer review training programs such as *Peerspectives*. For example, a total of 76 group review reports, each containing input from four participants and an editor-mentor, were submitted to the partnering journal and used to inform its editorial decision-making process. Not only did this format allow students to tangibly contribute to actual peer review, but their contact details were also shared with *The BMJ* for potential invitations to review manuscripts in the future, thereby adding trained reviewers to its reviewer pool. Moreover, having spent less time in professional scientific hierarchies, ECRs may be less prone to some known conscious and unconscious reviewer biases (e.g., prestige, affiliation).[4,50] ECRs are particularly receptive to learning and adapting new research methods and tools and are usually interested and motivated to improve science and quality assurance efforts.[21,51]

At first glance, there appears to numerous existing efforts to train peer review-related knowledge and skills with the aim of improving review quality, especially in asynchronous, condensed online formats, developed, curated, and maintained by major publishers.[24–28,52,53] Researcher-initiated programs like *Peerspectives* may offer different perspectives on peer review and scientific publishing topics than publisher-initiated programs.

Most peer review training initiatives are not formally evaluated for effectiveness, and the interventions are highly heterogeneous. A recent Cochrane review found few randomized controlled trials evaluating journal peer review training interventions, and concluded that training journal peer reviewers may lead to little or no improvement in quality.[54] Most of the evidence synthesized was low certainty, and the authors emphasized the necessity for further research with larger participant groups and including a wider range of valid and reliable outcome measures.[54] Two earlier systematic reviews specific to health research found that the evidence base on whether training (broadly defined) improved peer review-related knowledge and quality was inconclusive while highlighting considerable gaps and potential biases in existing studies.[55,56] The reviews called for more rigorous intervention studies evaluating training for peer reviewers.[55,56]

We identified only a few existing studies similar to our initiative. One of these, an evaluation of the EMEUNET Peer Review Mentoring Program for young rheumatology researchers, recommended: (1) the promotion of ‘face-to-face’ meetings between mentor and mentees, (2) an introductory video-conference meeting, to discuss a ‘practice’ manuscript and literature on peer review, and (3) organizing webinars with experienced reviewers.[57] All of these suggested elements were also realized in the *Peerspectives* curriculum.

Our initiative was unique in having participants contribute to four actual reviews at a top-tier biomedical journal during the workshops, with the option to earn ECTS credits applicable to doctoral course requirements. Participants’ names appeared alongside the editor-mentor’s on group review reports, fostering recognition, accountability, and likely greater engagement compared to passive training programs disconnected from real review processes. The Peerspectives course required significant donated time and effort from instructors, coordinators, and editor-mentors. Peer review training is typically not part of standard curricula nor formally recognized or incentivized by institutions.[22] In an effort to improve the sustainability of peer review training initiatives, we are sharing all *Peerspectives* lecture materials and a train-the-trainer handbook, both available on our OSF project page (https://osf.io/wyegc/). The handbook includes communication and organization templates, a course timeline, and suggestions to simplify implementation for prospective instructors. With these efforts, we aim to promote the expansion of the initiative and facilitate the launch of spin-off courses at other institutions and with other journal partners.

### Strengths and limitations

Strengths of our study include an independent assessment of participants’ peer review reports for the primary outcome by two experienced *BMJ* editors who were blinded to whether the review report had been written before or after course completion. Since the assessments were based on scientific manuscripts contemporaneously undergoing peer review at *The BMJ* and conducted in a standard review time frame, we believe we successfully mirrored conditions encountered by actual peer reviewers. Moreover, the study participation rate was very high among doctoral students enrolling in the course; out of 82 total students, 80 consented and enrolled in the study, and 78 completed pre- and post-intervention assessments for the primary outcome.

The main limitation of our study is its pre-post design. As an elective course formally embedded in a structured PhD program for which students could receive credit, our primary obligation was to provide training, with the scientific evaluation study being secondary. Consequently, it was not feasible to include a control group of study participants who did not participate in the course. Our ability to draw causal conclusions therefore relies on the assumption of stationarity over time of the outcomes that would have been measured if no intervention had been implemented (untreated counterfactuals).[36,37] Essentially, we relied on the assumption that no change would have occurred in the primary outcome between pre- and post-assessments if the study participants had not taken part in the *Peerspectives* course in-between the measurements. Although time-invariant determinants of the outcome are accounted for by design,[37,58] the presence of time-varying factors influencing the outcome may have introduced some bias in our results.[35–37,58] For example, we cannot exclude that if participants had not taken *Peerspectives*, they may have engaged in other activities during the semester that would have improved their peer review-related knowledge and skills (e.g., other PhD courses or their own research activities).

This potential “natural” improvement of students’ scores over time is a known issue when evaluating education intervention using pre-post studies[36]. However, given the short time frame between assessments (approximately 4.5 months) and the specificity of peer review-related knowledge and skills, it is unlikely that these external factors can explain the observed improvements. Indeed, we found that peer review quality assessed before the intervention was very weakly associated with age (increased on average by 0.02 for a 1-year increase in age) and was not associated with the number of prior reviews, suggesting that participants’ natural improvement over time is negligible (**Supplementary Material D**). We only looked at short-term outcomes, and we did not investigate whether the effect on quality is sustained in the longer-term.

Furthermore, we cannot fully ensure that the manuscripts used in the pre- and post-intervention assessments were of similar difficulty to review. We attempted to mitigate this issue by requesting four manuscripts of similar complexity from *BMJ* editors each semester. The similarity in median RQI scores by the handling editors across all manuscripts strengthens our confidence in their comparability (**Supplementary Material E**). Moreover, by randomizing the sequence of assigned manuscripts, we aimed at making the difficulty of the pre- and post-assessment the same, on average.

Another limitation is our use of the global RQI score as assessed by *BMJ* editors. While *The BMJ* editors routinely use this overall score for internal routine reviewer evaluation, a single-item global score for review quality does not necessarily capture the multi-dimensionality of this construct; single-item scales are also less reliable than multi-item scales.[59] Moderate inter-rater agreement in scores suggests that editors may weigh quality-related aspects of peer reviews differently when using the single-item global score. In an attempt to address this issue, we used the average of the RQI global scores from two different raters as the primary outcome. Lastly, in the secondary analysis comparing course participants to *BMJ* reviewers, we used only the handling editor’s RQI scores, as they were the sole evaluators of the actual *BMJ* reviewers’ reports. While this ensured consistency in the scoring for each manuscript, the handling editor was not blinded to the identity of the article’s assigned *BMJ* reviewers (due to open peer review) and they knew whether the report had been produced by *Peerspectives* participants and not actual *BMJ* reviewers. Additionally, *The BMJ* reviewers’ scores were assigned in real-time when the paper was under consideration at the journal, whereas Peerspectives participants’ reviews were scored all at once; up to two years later.

## CONCLUSIONS

Our findings provide evidence that participation in the *Peerspectives* program meaningfully improved the *BMJ* editor-judged quality of peer review reports drafted by doctoral students. However, the peer review reports generated after participating in the course were not non-inferior to those submitted by invited *BMJ* reviewers, who generally have more advanced career stages and reviewing experience. We observed considerable improvements in self-assessed knowledge and relevant peer review competencies following course participation.

## SUMMARY BOXES

### What is already known on this topic

● Editors often face difficulties in recruiting peer reviewers who can provide high-quality reviews within tight deadlines.
● Peer review is an essential cornerstone of the current publishing system but is rarely included in advanced academic curricula.
● While various reviewer training materials exist, few studies have assessed the effect of peer review training on review quality, especially among early career researchers.

### What this study adds

● Participation in a semester-long peer review training program meaningfully improved the quality of peer review reports drafted by doctoral students.
● However, these reports were not non-inferior to those submitted by invited BMJ reviewers, who generally have more advanced career stages and reviewing experience.
● Self-assessed knowledge and relevant peer review competencies improved substantially following course participation.

## ETHICS STATEMENTS

### Ethical approval

This study was approved by the Charité – Universitätsmedizin Berlin ethics committee (EA4/190/20). All participants provided written informed consent.

## DATA AVAILABILITY STATEMENT

The corresponding pseudonymised datasets used to generate our results and corresponding analytical code will be made accessible via our OSF project page upon publication: https://osf.io/wyegc/. Together with the study results, a train-the-trainer handbook and customizable materials will also be made available to facilitate course roll-out at other interested institutions.

## Supporting information

STROBE checklist

## Data Availability

The corresponding pseudonymised datasets used to generate our results and corresponding analytical code will be made accessible via our OSF project page upon publication.

https://osf.io/wyegc/

## Acknowledgements

They say “it takes a village”; indeed, without many motivated contributors, *Peerspectives* would not have been possible. First, we thank our editor-mentors who invested considerable time and energy to help train students to be critical readers and peer reviewers of the future. During the study period, editor-mentors included: Susanne Breitner-Busch, Ralph Brinks, Kristina Fišter, Lars Hemkens, André Karch, Hans-Georg Kuhn, Mariska Leeflang, Rutger Middelburg, Maarten van Smeden, and Bob Siegerink. We are grateful to Antoinette Maassen van den Brink for providing content about reporting sex and gender aspects. We thank Bob Siegerink for initiating the BIH SPOKES Wellcome Trust Funded Translational Partnership Fellowship program. We are extremely grateful to the program’s organizers, Tracey Weissgerber and Constance Holman, and our cohort of fellows for their valuable insights, support, and encouragement during the formative stages of this project. Finally, we are grateful to Tim Feeney, Elizabeth Loder, Tiago Villanueva, and Wim Weber, research editors at *The BMJ*, who were involved in selecting suitable manuscripts for the pre- and post-course assessments and scoring participants’ review reports.

## Contributors

JLR, SS, MP, TK, and TG were involved in study conception and design. JLR, NW, HG, IA, MLS, and TG were involved in data collection, curation, and course administration. KR and TG performed the statistical analyses. JLR and TG wrote the first draft of the manuscript, which was subsequently revised and approved by all authors. JLR and TG are the guarantors. The corresponding author attests that all listed authors meet authorship criteria and that no others meeting the criteria have been omitted.

## Transparency

JLR and TG (the manuscript’s guarantors) affirm that the manuscript is an honest, accurate, and transparent account of the study being reported; that no important aspects of the study have been omitted; and that any discrepancies from the study as originally planned (and, if relevant, registered) have been explained.

## Dissemination To Participants And Related Patient And Public Communities

In addition to the publication of our study findings in an international peer reviewed journal, we have presented our study design and course format at two large international conferences and a series of smaller scientific meetings. We have and will continue to make related information available on social media and our program’s website. All study participants have received a summary of our research study findings together with a link to the preprint. Upon publication, we plan to make all materials needed to deliver the course publicly available on our OSF project page: https://osf.io/wyegc/.

## REFERENCES

1 Ross-Hellauer T. What is open peer review? A systematic review. F1000Res. 2017;6:588. doi: 10.12688/f1000research.11369.2

2 Horbach SPJMS, Halffman WW. The changing forms and expectations of peer review. Res Integr Peer Rev. 2018;3:8. doi: 10.1186/s41073-018-0051-5

3 Allen H, Cury A, Gaston T, et al. What does better peer review look like? Underlying principles and recommendations for better practice: What does better peer review look like? Learn Publ. 2019;32:163–75. doi: 10.1002/leap.1222

4 Tennant JP, Ross-Hellauer T. The limitations to our understanding of peer review. Res Integr Peer Rev. 2020;5:6. doi: 10.1186/s41073-020-00092-1

5 Severin A, Chataway J. Overburdening of peer reviewers: A multi-stakeholder perspective on causes and effects. Learn Publ. 2021;34:537–46. doi: 10.1002/leap.1392

6 Dance A. Stop the peer-review treadmill. I want to get off. Nature. 2023;614:581–3. doi: 10.1038/d41586-023-00403-8

7 Pontille D, Torny D. From Manuscript Evaluation to Article Valuation: The Changing Technologies of Journal Peer Review. Hum Stud. 2015;38:57–79. doi: 10.1007/s10746-014-9335-z

8 Tennant JP, Dugan JM, Graziotin D, et al. A multi-disciplinary perspective on emergent and future innovations in peer review. F1000Res. 2017;6:1151. doi: 10.12688/f1000research.12037.3

9 Waltman L, Kaltenbrunner W, Pinfield S, et al. How to improve scientific peer review: Four schools of thought. Learn Publ. 2023;36:334–47. doi: 10.1002/leap.1544

10 Publons, Publons. Publons’ Global State Of Peer Review 2018. 2018.

11 Ho RC-M, Mak K-K, Tao R, et al. Views on the peer review system of biomedical journals: an online survey of academics from high-ranking universities. BMC Med Res Methodol. 2013;13:74. doi: 10.1186/1471-2288-13-74

12 Warne V. Rewarding reviewers - sense or sensibility? A Wiley study explained: Rewarding reviewers - sense or sensibility? Learn Publ. 2016;29:41–50. doi: 10.1002/leap.1002

13 Willis JV, Ramos J, Cobey KD, et al. Knowledge and motivations of training in peer review: An international cross-sectional survey. PLoS One. 2023;18:e0287660. doi: 10.1371/journal.pone.0287660

14 Mulligan A, Hall L, Raphael E. Peer review in a changing world: An international study measuring the attitudes of researchers. J Am Soc Inf Sci Technol. 2013;64:132–61. doi: 10.1002/asi.22798

15 Heinemann L. Reviewer: an endangered species?! J Diabetes Sci Technol. 2015;9:167–8. doi: 10.1177/1932296814563883

16 Aczel B, Szaszi B, Holcombe AO. A billion-dollar donation: estimating the cost of researchers’ time spent on peer review. Res Integr Peer Rev. 2021;6:14. doi: 10.1186/s41073-021-00118-2

17 Aczel B, Barwich A-S, Diekman AB, et al. The present and future of peer review: Ideas, interventions, and evidence. Proc Natl Acad Sci U S A. 2025;122:e2401232121. doi: 10.1073/pnas.2401232121

18 Hanson MA, Barreiro PG, Crosetto P, et al. The strain on scientific publishing. arXiv [cs.DL]. 2023.

19 Kurth T, Piccininni M, Loder EW, et al. Parallel pandemic: The crush of covid-19 publications tests the capacity of scientific publishing. BMJ Opinion. Published Online First: 2020.

20 Björk B-C, Solomon D. The publishing delay in scholarly peer-reviewed journals. J Informetr. 2013;7:914–23. doi: 10.1016/j.joi.2013.09.001

21 Kent BA, Holman C, Amoako E, et al. Recommendations for empowering early career researchers to improve research culture and practice. PLoS Biol. 2022;20:e3001680. doi: 10.1371/journal.pbio.3001680

22 Kusumoto FM, Bittl JA, Creager MA, et al. Challenges and controversies in peer review: JACC review topic of the week. J Am Coll Cardiol. 2023;82:2054–62. doi: 10.1016/j.jacc.2023.08.056

23 EQUATOR network. Peer review training and resources. 2021. https://www.equator-network.org/toolkits/peer-reviewing-research/peer-review-training-and-r esources/#PRTraining (accessed 4 March 2021)

24 Focus on Peer Review. https://masterclasses.nature.com/online-course-on-peer-review/16507836 (accessed 6 May 2024)

25 Training peer reviewers. J Food Sci. 2023;88:3621–2. doi: 10.1111/1750-3841.16758

26 Certified Peer Reviewer course. https://researcheracademy.elsevier.com/navigating-peer-review/certified-peer-reviewer-cour se (accessed 6 May 2024)

27 Peer reviewer training course. https://wkauthorservices.editage.com/peer-reviewer-training-course/ (accessed 6 May 2024)

28 Excellence in Peer Review: Reviewer training network. Editor Resources. 2021. https://editorresources.taylorandfrancis.com/reviewer-guidelines/peer-review-training/ (accessed 6 May 2024)

29 Xu J, Kim K, Kurtz M, et al. Mentored peer reviewing for PhD faculty and students. Nurse Educ Today. 2016;37:1–2. doi: 10.1016/j.nedt.2015.11.031

30 Podder V, Price A, Sivapuram MS, et al. Collective Conversational Peer Review of Journal Submission: A Tool to Integrate Medical Education and Practice. Ann Neurosci. 2018;25:112–9. doi: 10.1159/000488135

31 Schroter S, Black N, Evans S, et al. Effects of training on quality of peer review: randomised controlled trial. BMJ. 2004;328:673.

32 Callaham ML, Knopp RK, Gallagher EJ. Effect of written feedback by editors on quality of reviews: two randomized trials. JAMA. 2002;287:2781–3. doi: 10.1001/jama.287.21.2781

33 Houry D, Green S, Callaham M. Does mentoring new peer reviewers improve review quality? A randomized trial. BMC Med Educ. 2012;12:83. doi: 10.1186/1472-6920-12-83

34 Wong VSS, Strowd RE 3rd, Aragón-García R, et al. Mentored peer review of standardized manuscripts as a teaching tool for residents: a pilot randomized controlled multi-center study. Res Integr Peer Rev. 2017;2:6. doi: 10.1186/s41073-017-0032-0

35 Aggarwal R, Ranganathan P. Study designs: Part 4 - Interventional studies. Perspect Clin Res. 2019;10:137–9. doi: 10.4103/picr.PICR_91_19

36 Stuart EA. Estimating causal effects using school-level data sets. Educ Res. 2007;36:187–98. doi: 10.3102/0013189x07303396

37 Piccininni M, Stensrud MJ, Shahn Z, et al. Causal inference for N-of-1 trials. arXiv [stat.ME]. 2024.

38 Naydenov A. PaperHive – a coworking hub for researchers that aims to make reading more collaborative. Impact of Social Sciences Blog. Published Online First: 15 June 2016.

39 European Commission: Directorate-General for Education, Youth, Culture SA. *ECTS users’ guide* 2015. Publications Office of the European Union 2015.

40 Coppock A. Easy-to-Use Tools for Common Forms of Random Assignment and Sampling [R package randomizr version 0.24.0]. Published Online First: 10 February 2023.

41 van Rooyen S, Black N, Godlee F. Development of the review quality instrument (RQI) for assessing peer reviews of manuscripts. J Clin Epidemiol. 1999;52:625–9. doi: 10.1016/s0895-4356(99)00047-5

42 Wong VSS, Strowd RE, Aragón-García R, et al. Mentored peer review of standardized manuscripts as a teaching tool for residents: a pilot randomized controlled multi-center study. Research Integrity and Peer Review. 2017;2.

43 Shrauger JS, Schohn M. Self-Confidence in College Students: Conceptualization, Measurement, and Behavioral Implications. Assessment. 1995;2:255–78. doi: 10.1177/1073191195002003006

44 Cui Y, Konietschke F, Harrar SW. The nonparametric Behrens-Fisher problem in partially complete clustered data. Biom J. 2021;63:148–67. doi: 10.1002/bimj.201900310

45 Brunner E, Munzel U. The nonparametric Behrens-fisher problem: Asymptotic theory and a small-sample approximation. Biom J. 2000;42:17–25. doi: 10.1002/(sici)1521-4036(200001)42:1<17::aid-bimj17>3.0.co;2-u

46 Revelle W. psych: Procedures for Personality and Psychological Research. Evanston, Illinois, USA: Northwestern University, 2022.

47 van Rooyen S, Godlee F, Evans S, et al. Effect of open peer review on quality of reviews and on reviewers’ recommendations: a randomised trial. BMJ. 1999;318:23–7. doi: 10.1136/bmj.318.7175.23

48 Vollmar M. Neue Promovierendenstatistik: Analyse der ersten Erhebung 2017. Wirtsch Stat.

49 The scarce peer reviewer and challenges journal editors face. Editage Insights. 2015. https://www.editage.com/insights/the-scarce-peer-reviewer-and-challenges-journal-editors-face (accessed 2 November 2024)

50 Lee CJ, Sugimoto CR, Zhang G, et al. Bias in peer review. J Am Soc Inf Sci Technol. 2013;64:2–17. doi: 10.1002/asi.22784

51 Campbell HA, Micheli-Campbell MA, Udyawer V. Early Career Researchers Embrace Data Sharing. Trends Ecol Evol. 2019;34:95–8. doi: 10.1016/j.tree.2018.11.010

52. 52 Reviewing in the Sciences. https://webofscienceacademy.clarivate.com/learn/courses/128/Reviewing%20in%20the%20 Sciences (accessed 13 December 2024)

53. 53 An Introduction to Peer Review. https://webofscienceacademy.clarivate.com/learn/courses/119/an-introduction-to-peer-revie w (accessed 13 December 2024)

54 Hesselberg J-O, Dalsbø TK, Stromme H, et al. Reviewer training for improving grant and journal peer review. Cochrane Database Syst Rev. 2023;11:MR000056. doi: 10.1002/14651858.MR000056.pub2

55 Galipeau J, Moher D, Campbell C, et al. A systematic review highlights a knowledge gap regarding the effectiveness of health-related training programs in journalology. J Clin Epidemiol. 2015;68:257–65. doi: 10.1016/j.jclinepi.2014.09.024

56 Bruce R, Chauvin A, Trinquart L, et al. Impact of interventions to improve the quality of peer review of biomedical journals: a systematic review and meta-analysis. BMC Med. 2016;14:85. doi: 10.1186/s12916-016-0631-5

57 Rodríguez-Carrio J, Putrik P, Sepriano A, et al. Improving the peer review skills of young rheumatologists and researchers in rheumatology: the EMEUNET Peer Review Mentoring Program. RMD Open. 2018;4:e000619. doi: 10.1136/rmdopen-2017-000619

58 Nianogo RA, Benmarhnia T, O’Neill S. A comparison of quasi-experimental methods with data before and after an intervention: an introduction for epidemiologists and a simulation study. Int J Epidemiol. 2023;52:1522–33. doi: 10.1093/ije/dyad032

59 Nunnally JC, Bernstein I. Psychometric Theory. 3rd ed. Maidenhead, England: McGraw Hill Higher Education 1994.

