## Supplementary Materials for "Engaging doctoral students in peer review: a pre-post study evaluating the effectiveness of the “Peerspectives” course on review quality, knowledge and skills"

**Contents:**


**Supplementary Material A. Personal details questionnaire 2**

**Supplementary Material B. Sample course syllabus 3**

**Supplementary Material C. Self-assessment of peer review-related knowledge and abilities 8**

**Supplementary Material D. Pre-intervention Review Quality Instrument scores by age and prior reviewing experience 9**

**Supplementary Material E. Median Review Quality Instrument scores per manuscript 10**

**Supplementary Material F. Selection of anonymous responses to the course evaluation question about whether participants would recommend the course and to which target audience 11**

### Supplementary Material A. Personal details questionnaire

1. Have you ever performed a peer review of a scientific manuscript in the past? (yes / no)
2. If so, how many? (free response, integer $\geq 0$ expected)
3. Age (based on birthday in MM/YY format)
4. Gender (Male / Female / Other / Prefer not to say)
5. Information about current educational level (i.e. Master's student, doctoral/PhD student or postdoc), type of program and expected graduation date (MM/YY)
6. Prior participation in a epidemiological / biostatistical methods-focused course (yes / no)
7. Prior self-training in biostatistics/epidemiology (yes / no)

### Supplementary Material B. Sample course syllabus

peerspectives

**Peer review and biomedical editing training initiative**

**Course Syllabus - Sample Semester**
*in collaboration with a partnering journal (The BMJ) and international experts*

#### Course Structure and Contacts

| | |
|---|---|
| Format | Interactive lectures and workshops |
| Dates | Between April and July, on selected Fridays (some workshop dates may differ by group) |
| Times | Lectures 13:00-16:00 CEST<br>Workshops may vary by group (*See dates at the end of the syllabus. Let us know about potential conflicts early on so we can try to accommodate you when assigning groups.) |
| Location | Online - Zoom |
| Credits | 4 ECTS points for full attendance and successful completion of all assignments and contributions to four group peer review reports |

#### Purpose

To provide modern training and insights into the structure, purpose, and conduct of the peer-review and editing processes in journals with a general biomedical and/or population health focus. The course will include a series of instructive lectures and mentor-guided small group workshops providing hands-on experience in reviewing and editing submitted research papers from participating scientific journals.

#### Rationale

Scientific journals in the biomedical sciences publish scholarly articles and provide an important platform for transparent presentation, exchange, and discussion of new scientific developments. Peer review, in all its forms, plays an integral role in ensuring the integrity and quality of the scholarly record of journals. Despite this, curricula of advanced academic programs generally lack formal training in the structure, purpose, and conduct of peer review and editing. As a consequence, the first peer reviews of early career researchers are often conducted in a self-guided, learning-by-doing setting without formal training or feedback. This not only potentially leads to a slowed peer-review process and possibly an incorrect interpretation of the role of a peer reviewer, but can also lead to unhelpful or even unusable reviews for editors and authors of submitted manuscripts alike. Furthermore, many journals find it difficult to find qualified individuals willing to take on review assignments and submit high-quality reviews in a timely manner. The Peerspectives program aims to address these needs with an innovative, mentor-guided, mutually beneficial training program.

#### Learning objectives

By providing formal training through a hybrid structure of interactive lectures and hands-on workshops, upon successful completion of the training program, Peerspectives participants will:

1. have an increased familiarity with modern statistical and epidemiological methods for biomedical research
2. have an improved understanding of elements of study designs encountered in the biomedical literature
3. gain familiarity with reporting guidelines for scientific manuscripts, with emphasis on the link between objectives, suitable methods, reporting of results, and appropriate interpretation
4. learn how to detect common biases and fallacies encountered in the medical literature, where to look for them, and best practices to minimize them in study design and analysis
5. better understand the role of journals, editors, and peer reviewers within the scientific process
6. practice and understand the value of clear communication and efficiency in the review process
7. hone critical thinking and assessment skills
8. practice giving constructive and helpful scientific critique
9. learn the importance of transparency and adherence to ethical and methodological guidelines
10. become acquainted with open-access, open review concepts, pre-prints
11. develop an understanding of what reviewers and editors are looking for in scientific writing and improve the quality of their own future manuscripts
12. learn the importance of, and ways to, consider sex- and gender-related aspects in scientific research and in the context of peer review

#### Peerspectives approach

The Peerspectives program is a semester-long program with two main blocks: interactive lectures and hands-on practical workshops. The following steps describe our approach in detail:

1. The program starts with **four, 3-hour interactive lectures** (topics outlined below). Each lecture will be supplemented by a homework assignment, which must be completed before the next lecture.

2. After the successful completion of the lectures, small groups of 4 trainees will be paired with an experienced editor-mentor for a series of four workshops.
    a. A journal editor will select a suitable manuscript that would benefit from a methods-focused review from the journal's recent submissions and assign it to the editor-mentor
    b. The PDF of the manuscript and any supplemental materials will be uploaded to a specifically prepared, secured channel in PaperHive (https://paperhive.org/) to promote transparent and real-time discussions of the manuscript by the trainees and the mentor.
    c. A review report will then be drafted together by the trainees in a "living" shared document. A different trainee will be "in charge" of each manuscript (on a rotating basis). This leader will be responsible for gathering and consolidating the group's feedback and finalizing the first draft. This draft will then be revised and discussed together with the mentor during the workshop.
    d. Once finalized, the review report will be submitted to the participating journal by the mentor. The names of the four trainees involved in the review will be included in the review report.
    e. The decision by the journal and other reviewers' comments will be forwarded to the group members and discussed in future workshop sessions (when possible).

**Assessment**

To receive credit for the course, you are required to submit two peer review reports, once before the course and once at the end of the course (pre- and post-intervention assessments). For these assessments, you are required to draft a peer review report *on your own* under simulated real-world conditions. These assessments will also be used for the linked scientific evaluation study, in which your participation is invited but entirely optional. These assessments will not be graded, but will help the instructor team tailor the course to your needs, assess your progress, and optimize the course for future runs. You can also use this experience to assess your own confidence in reviewing both before and after participating in the program. Your choice to participate in the study (or not) will have no impact on your ability to successfully complete the course.

The pre- assessment is due **on APRIL DATE by 12:00 noon** and the post-assessment on **AUGUST DATE by 12:00 noon.** Please submit it as a PDF file attachment via reply email with the subject line "Peerspectives assessment: *YOUR LAST NAME***"**.

**Paperhive**

We will use the tool Paperhive to facilitate communication between workshop group members about the manuscripts during the group review process in the week prior to each workshop. These live manuscripts shared on Paperhive must be kept strictly confidential, as you are bound to the conditions of the confidentiality agreement you signed at the beginning of the course. To use Paperhive, please first create an account here: https://paperhive.org/. You can use any email address to register. Then, once logged in, you should join your unique workshop group's channel using the invitation link you will receive by email.

**Schedule**

Notes:

1. An optional “listening in” session for one of the editorial manuscript meetings will be scheduled at the end of the course.
2. Since we have a limited number of sessions and given the nature of the mentor-led group workshops, attendance is mandatory for all sessions of the program. We encourage participants to mark their calendars with all dates as soon as they are registered for the course. If you know in advance you cannot make a particular session, please notify us as soon as possible so we can make necessary arrangements.
3. You will need to exchange with your workshop group outside of class hours to prepare a first draft of the group’s peer review report *before* each workshop session. Be sure to budget some time for this in your schedule!

| Date & Time | Topic | Content | Assignment / outside of class work |
|---|---|---|---|
| Due via email by end of March date | Course enrollment<br><br>Optional: study participation | Sign confidentiality agreement<br>Optional: informed consent form, self-assessment survey, personal details form | Please submit the requested documentation to us via email |
| Assigned end of March<br><br>Due via email by mid-April date | Assessment 1: Pre-course peer review report | Full details will be sent via email; please read instructions carefully | Submit via reply email with subject line: “Peerspectives pre-course assessment: *YOURLASTNAME*” |
| Mid-April date, 3 hours | Lecture 1 | Course overview & structure<br>The role of academic/scientific journals, the role of editors, the role of authors | Take-home assignment #1: Finding suitable peer reviewers. Due via email by day before next class at noon |
| Late April date, 3 hours | Lecture 2 | Ethical guidelines for peer review (COPE)<br>Open science<br>Sex and gender aspects in peer review | Take-home assignment #2: Published paper with open data / code - reproducibility and reporting<br>Submit via online form by day before next class at noon |
| Early May date, 3 hours | Lecture 3 | How to conduct a “good” peer review<br>Methodological guidelines for peer review; elements of a good peer review report & writing tips (focused on clarity, quality, efficiency) | Take-home assignment #3: Critique of two peer review reports<br>Submit via online form by day before next class at noon |

| | | | |
|---|---|---|---|
| Mid-May date,<br>3 hours | Lecture 4 | From start to finish: how to review a paper<br>Expectations- first "live" review assignment, workshop logistics<br>Wrap up | Assignment: As a group, generate first draft of peer review report for 1st under-review manuscript (to be finalized and submitted at the end of Workshop 1) |
| Date in early June,<br>3 hours | Workshop 1 | Finalize first 'live' peer review & submit via editor-mentor | Assignment: As a group, generate first draft of review report for 2nd under-review manuscript (To be finalized at end of Workshop 2) |
| Date in mid-June,<br>3 hours | Workshop 2 | Finalize 2nd 'live' peer review & submit via editor-mentor | Assignment: As a group, generate first draft of review report for 3rd under-review manuscript (To be finalized at end of Workshop 3) |
| Date in early July,<br>3 hours | Workshop 3 | Finalize 3rd 'live' peer review & submit via editor-mentor | Assignment: As a group, generate first draft of review report for 4th under-review manuscript (To be finalized at end of Workshop 4) |
| Date in mid-July,<br>3 hours | Workshop 4 | Finalize 4th 'live' peer review & submit via editor-mentor | Complete post-course peer review report assessment<br>(see next item) |
| Assigned after Workshop 4<br><br>Due via email by mid-August date | Assessment 2: Post-course peer review report | Full details will be sent after Workshop 4 via email; please read instructions carefully<br>Course evaluation<br>(Optional: Final self-assessment survey: study participants only) | Submit via email with subject line: "Peerspectives Post-course Assessment: *YOURLASTNAME*" |
| Late August,<br>3 hours | Optional: Wrap-up and listening-in session | Listening in on editorial manuscript meeting with partner journal followed by reflection and course wrap up discussion | -- |

### Supplementary Material C. Self-assessment of peer review-related knowledge and abilities

Questionnaire to assess the self-perceived confidence in peer review-related knowledge and abilities (Answer options 1 to 5; 1 = strongly disagree to 5 = strongly agree). Administered as an online survey to participants once prior to Peerspectives course (baseline self-assessment) and once after the course (final self-assessment). Modified from Shrauger and Schohn.[43]

**Question**

1. I feel confident in my ability to peer review scientific manuscripts.
2. I have a clear understanding of what is expected from a reviewer.
3. I know what to look for in a manuscript when conducting a peer review.
4. I feel confident and comfortable using reporting guidelines.
5. I have a clear understanding of ethical aspects relevant in peer review.
6. I feel confident in formulating constructive and clear scientific critique.
7. I feel confident in my own scientific writing abilities.
8. I am aware of key sex- and gender-related considerations relevant for peer review.

### Supplementary Material D. Pre-intervention Review Quality Instrument scores by age and prior reviewing experience

To visualize how pre-intervention scores differed by age and prior review experience, we created two dot plots and overlaid two univariate linear regression models fitted with the pre-intervention primary outcome (mean global Review Quality Instrument (RQI) score) as the dependent variable and age and number of prior reviews as independent variable. We observed a negligible increase in RQI for each year increase in age (0.02, 95% CI: [-0.02, 0.05]) but not for the number of prior reviews (0.00, 95% CI: [-0.06, 0.06]).

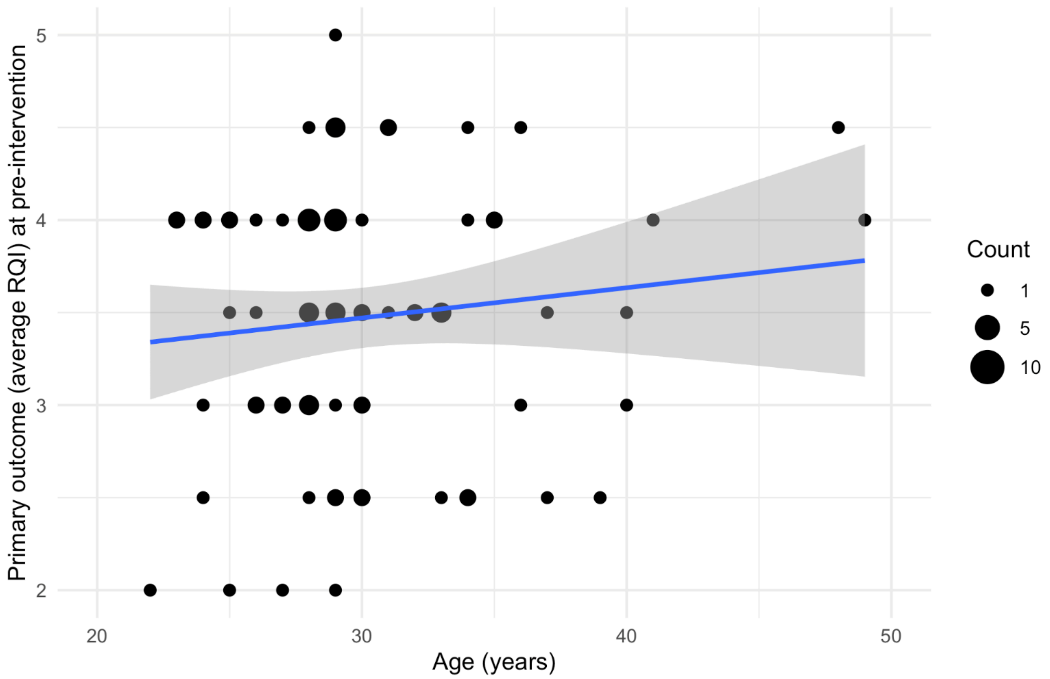


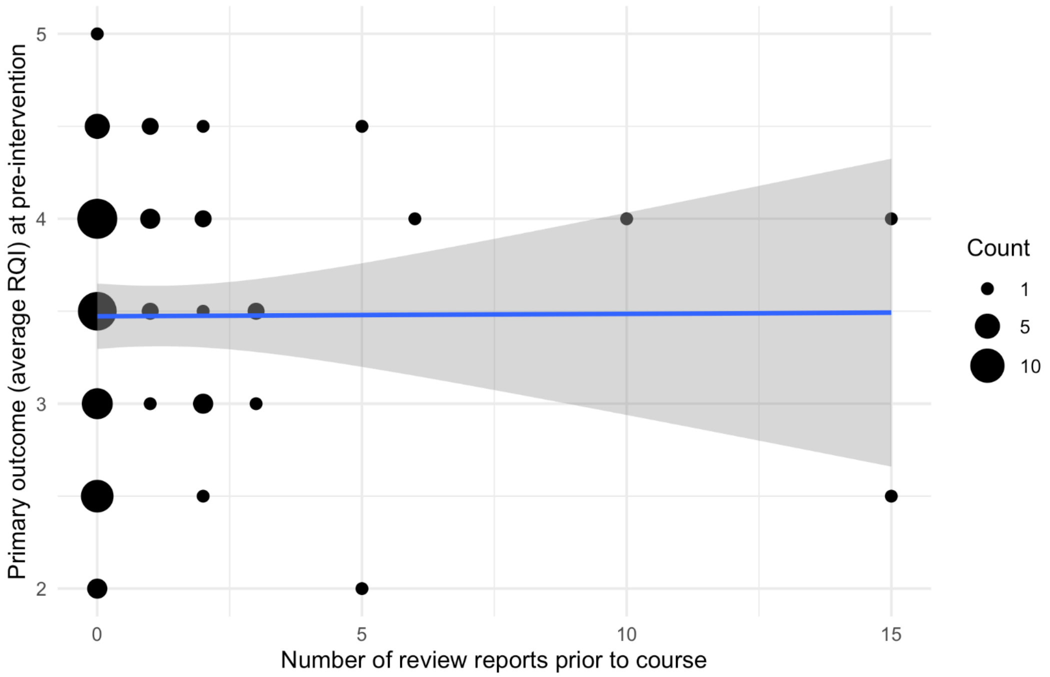

### Supplementary Material E. Median Review Quality Instrument scores per manuscript

Median RQI global score per manuscript as judged by the handling editor for participants who attended Peerspectives and actual BMJ reviewers. N denotes the number of reviews performed for each manuscript.

| **Manuscript** | **Cohort** | **Post-Course Participants** Median RQI (N) | ***BMJ* Reviewers** Median RQI (N) |
|---|---|---|---|
| 1 | Fall 2020 | 4.0 (4) | 4.0 (3) |
| 2 | Fall 2020 | 3.5 (4) | 4.0 (4) |
| 3 | Fall 2020 | 4.0 (4) | 5.0 (3) |
| 4 | Fall 2020 | 4.0 (4) | 4.0 (2) |
| 5 | Spring 2021 | 4.0 (4) | 4.0 (3) |
| 6 | Spring 2021 | 4.0 (5) | 3.5 (2) |
| 7 | Spring 2021 | 4.0 (5) | 4.5 (4) |
| 8 | Spring 2021 | 3.5 (6) | 4.5 (2) |
| 9 | Fall 2021 | 3.0 (6) | 4.0 (5) |
| 10 | Fall 2021 | 4.0 (5) | 4.0 (3) |
| 11 | Fall 2021 | 3.5 (4) | 4.0 (3) |
| 12 | Fall 2021 | 3.5 (6) | 4.0 (2) |
| 13 | Spring 2022 | 4.0 (6) | 5.0 (3) |
| 14 | Spring 2022 | 4.0 (5) | 4.5 (2) |
| 15 | Spring 2022 | 3.0 (5) | 4.0 (4) |
| 16 | Spring 2022 | 4.0 (5) | 3.0 (2) |

### Supplementary Material F. Selection of anonymous responses to the course evaluation question about whether participants would recommend the course and to which target audience

- *“I would totally recommend the course to all early career researchers, as it gives a very good and clear introduction in how to do peer reviews. Even for researchers with a little more peer review experience, the practical aspects of the course might still be of interest.”*
- *“[This was] one of the courses which I benefited from the most with regard to future tasks in the field of research.”*
- *“I wish I had this course in my master's studies. I also wish that universities can offer more courses like this one, where students can learn the theory and then have a real-world supervised practical part. It is really a good example of how teaching in science should be.”*
- *“[Peerspectives] could be useful for literally anyone in the sciences.”*
- *“Peerspectives was one of the best courses I have ever attended. In science, it is so common to find yourself doing a complicated and important task without ever getting proper training for it. […] for the participants of this course, writing peer reviews will not be one of them.”*
- *“The course might be relevant for most PhD students everywhere. It's particularly relevant for PhD students with a focus on methodology, since they are often asked to perform peer review in a very early career stage.”*
- *“For me peer review was somewhat mysterious before the course, but now having had this course I gained a very clear understanding of what it is about. I especially appreciate the opportunity to practice the peer review process in a safe space.”*
- *“One of the most important classes I had during my PhD. Although the papers are quite hard, critical thinking is developed. One can not start early enough doing these types of reviews.”*
- *“I now feel more comfortable reviewing a manuscript. I used to sometimes feel lost in the manuscript, but now I can do a comprehensive review thoroughly, accurately and without getting lost in the details. The course has also helped me a lot with the language that should be used in peer review.”*
- *“I really liked our mentor and his insightful real life stories from reviewing papers. The interaction and coworking with the other team members was friendly, productive and straightforward.”*
- *“I feel so much more confident in my clinical research.”*
- *“This is a fantastic course. I am happy I was part of it. That it contributes to actual work for The BMJ and has an evaluation embedded makes it even better.”*
